# A Spatial Comparison of Molecular Features Associated with Resistance to Pembrolizumab in BCG Unresponsive Bladder Cancer

**DOI:** 10.1101/2023.11.28.23299093

**Authors:** Khyati Meghani, Noah Frydenlund, Yanni Yu, Bonnie Choy, Joshua J. Meeks

**Author notes:** Corresponding author:* Joshua J. Meeks,; @JoshMeeks, C (312)363-8959. Equal contribution (Khyati Meghani, Noah Frydenlund). **Take Home Message:** We conducted a spatial transcriptomic investigation of BCG unresponsive bladder cancers treated with pembrolizumab and identified distinct expression signatures of the tumor epithelium and tumor microenvironment associated with response or resistance that may be applied in the future to predict response to checkpoint immunotherapy.

## Abstract

**Background:** Intravenous immune checkpoint inhibition achieves a 40% three-month response in BCG-unresponsive non-muscle invasive bladder cancer (NMIBC) with carcinoma in situ (CIS). Yet only half of early responders will continue to be disease free by 12 months, and resistance mechanisms are poorly defined.

**Objective:** We assessed the molecular features associated with response to immunotherapy in BCG unresponsive non-muscle invasive bladder cancers treated with pembrolizumab.

**Design, Setting, and Participants:** We performed digital spatial profiling (DSP) of BCG unresponsive NMIBC tumors before and after IV pembrolizumab therapy.

**Intervention:** Pembrolizumab was administered intravenously in patients with NMIBC at the time of recurrence after BCG therapy. Biopsies were obtained before starting IV pembrolizumab and three months post-treatment.

**Outcomes and Statistical Analysis:** Spatial gene expression profiling of the tumor niche pre- and post IV pembrolizumab.

**Results and Limitations:** We evaluated 119 regions of interest (ROIs) from five patients, which included 60 epithelial (PanCK+) and 59 stromal segments (PanCK-). ROIs from responders had distinct expression signatures from non-responders for both the tumor and TME. Responders were more likely to have a dynamic change in expression after pembrolizumab than non-responders. A major limitation of this study was the number of patients evaluated.

**Conclusion:** For the first time, we have identified distinct expression signatures associated with response and resistance to IV pembrolizumab in NMIBCs. Further research with more patients and alternative checkpoint inhibitors is essential to validate our findings.

**Patient Summary:** We identify the molecular features of tumors associated with response to pembrolizumab for patients with BCG unresponsive NMIBCs.

## Introduction

BCG is the primary treatment for high-grade non-muscle invasive bladder cancer (NMBIC). Yet, at least one-third of NMIBCs treated with BCG will not respond and progress to more advanced stages of bladder cancer. In 2020, following the results of KEYNOTE-057^1,2^, in which 40.6% of patients had a complete response at three months, pembrolizumab monotherapy was approved for use in patients with BCG unresponsive high-risk NMIBC. Unfortunately, response to pembrolizumab is not durable, and overall, 80% of treated patients will have recurred or progressed by 12 months.

Since FDA approval, pembrolizumab has become a mainstay in the treatment of BCG unresponsive bladder cancer. Yet, there is limited information on how to best identify patients who will benefit from this course of treatment. The identification of biomarkers that predict response to pembrolizumab could facilitate the selection of NMIBCs, and spare unresponsive patients the unnecessary toxicity associated with immune checkpoint treatment.

We have previously performed a multi-omics evaluation of a small Phase I trial of BCG unresponsive NMIBCs treated with BCG and intravesical pembrolizumab^3^. While this study was primarily focused on mechanisms of tumor response in the unique setting of intravesical administration of pembrolizumab and BCG, we were intrigued to compare this to IV pembrolizumab. To evaluate the response mechanisms of IV pembrolizumab, we performed digital spatial profiling of tumors from responders and non-responders before and after treatment. Our results describe the spatial transcriptomic differences in pre-treatment NMIBCs and offer initial insights to identify individuals who may respond to IV pembrolizumab.

## Methods

### Sample identification and collection

After obtaining institutional review board (IRB) approval, the Northwestern Medicine Enterprise Data Warehouse (EDW) was queried to identify patients with BCG unresponsive NMIBC who were treated with at least three cycles of intravenous pembrolizumab. Patients were selected for inclusion in the study if adequate pre- and post-treatment formalin fixed paraffin embedded (FFPE) bladder biopsies were available in our institutional tissue repository. We identified five patients (three responders and two non-responders, Supplementary Table 1) who met inclusion criteria. FFPE blocks were sectioned at a thickness of 5μm and mounted on slides for DSP analysis. Slides were reviewed by a genitourinary pathologist (BC) to confirm the adequacy of selected biopsies before proceeding with DSP preparations.

### Digital Spatial Profiling

Using methods previously described^4^, slides were deparaffinized by baking in a drying oven at 60°C for two hours, washing in xylene (3 x 5 min), 100% ethanol (2 x 5 min), 95% ethanol (2 x 5 min), and 1x PBS (1 x 1 min). Target retrieval was performed by placing the slides into a steamer containing DEPC-water heated to 99°C for 10 seconds followed by 1x Tris-EDTA for 20 minutes. Slides were then washed in 1x PBS (5 min) before incubating in 1μg/mL Proteinase K (Thermo Fisher, Waltham, MA) at 37°C for 20 minutes. Post-fixation was performed by washing in 10% neutral buffered formalin (NBF) for five minutes, followed by NBF stop buffer (0.1M tris, 0.1M glycine in DEPC-treated water, 2 x 5 min), followed by 1x PBS (1 x 5 min). RNA probe library *in situ* hybridization was performed using the NanoString GeoMX™ Whole Transcriptome Atlas (WTA) (NanoString, Seattle WA) which was diluted per manufacturer instructions to create hybridization solution. Hybridization solution was applied to each slide which then covered with Grace Bio-Labs Hybrislip™ (Bend, OR) hybridization covers. Slides were incubated in a hybridization oven at 37°C for 20 hours. Next, off target probes were removed by performing stringent washes (100% formamide in equal parts 4X-SSC) at 37°C (2 x 25 min) followed by 2X SSC wash at room temperature (2 x 2 min). Slides were then placed into a humidity chamber at room temperature and blocked using blocking buffer W (NanoString, Seattle, WA) for 30 minutes. To facilitate identification of the tumor and TME slides were next stained with immunofluorescent antibodies from the NanoString solid tumor TME morphology kit [Pan-CK for epithelial cells, CD45 for immune cells, and SYTO 83 for nuclear staining] (NanoString, Seattle, WA) for one hour.

DSP was then performed on prepared slides using the GeoMx digital spatial profiler (NanoString, Seattle, WA). After loading and scanning the slides onto the instrument a total of 60 regions of interest (ROIs) were manually selected for transcriptomic profiling. Each ROI was divided into two segments based on the presence of immunohistochemical morphologic markers, with PanCK+ staining areas analyzed as tumor segments and PanCK-segments analyzed as stroma. Photocleaved DNA oligonucleotides were aspirated from each segment separately and stored in an individual well in a 96-well plate. Per the default NanoString GeoMx™ protocol, Illumina i5 and i7 dual indexing primers were added to the oligonucleotide tags during PCR, uniquely indexing each segment. AMPure XP beads (Beckman Coulter, Indianapolis, IN) were used for PCR purification. Library concentration was measured using a Qubit fluorometer (Thermo Fisher, Waltham, MA) and quality was assessed using a Bioanalyzer (Agilent Technologies Inc., Santa Clara, CA). Sequencing was performed on an Illumina NovaSeqX (Illumina Inc., San Diego, CA). After sequencing, reads were trimmed, stitched, aligned, and de-duplicated. Fastq files were converted to DCC files using the GeoMx NGS Pipeline v2.3.3.10, which were then loaded onto the GeoMx instrument and converted into target counts for each segment. Raw counts were filtered based on two criteria : to remove targets detected below the limit of quantitation (LOQ), and to remove segments with fewer than 50 nuclei. Filtered raw counts were Q3 normalized for comparison across all segments and were used for further analysis.

### Bioinformatics and Data Visualization

All analyses were performed in R v4.2.3. Principal component analysis was performed using PCAtools v2.10.0. Heatmaps were generated using ComplexHeatmap v2.14.0. Differential expression analysis was performed using limma v3.54.2. Volcano plots were generated using ggplot2 v3.4.4. Pathway analysis was conducted using msigdb^5^ gene sets downloaded using msigdbR v7.5.1 package and analyzed using fgsea v1.24.0. Gene signatures for the intravesical cohort were generated by comparing PanCK and Stroma specific expression profiles between responders and non-responders. Similarly, cluster-specific gene signatures for the PURE01 cohort^6^ were generated by comparing each cluster to the rest. Enrichment of these gene sets within the current IV pembrolizumab cohort was tested using the GSEA function, and GSEA plots were generated using the gseaplot function in clusterProfiler v4.6.2. Exhaustion score was calculated as a mean expression of PDCD1, HAVCR2, LAG3, CTLA4, and TIGIT genes. Inflammation score was generated as a mean expression of genes within the msigdb Hallmark Interferon alpha and Interferon gamma response gene sets^5^. Immune deconvolution of the stromal segments was performed using SpatialDecon v1.8.0. Immune Infiltration score was generated as a sum of the different immune cell types (T/NK cells, B cells, myeloid cells, neutrophils, and mast cells) identified by immune deconvolution.

## Results

### Cohort

We have previously performed bulk RNA sequencing to identify expression signatures of CPI-treated tumors across multiple cohorts^6^. Yet, due to the limited size of the tumor epithelium in NMIBCs with CIS in which tumors are a few cell layers we hypothesized that bulk RNA-seq might lack the resolution required to dissect the granular details of the tumor/TME in this setting. Therefore, in this study, we performed spatial profiling of tumor sections before and after intravenous pembrolizumab to identify 1) pre-treatment features associated with response or resistance and 2) describe how IV pembrolizumab alter the tumor/TME interaction in BCG unresponsive NMIBCs. Five patients treated with IV pembrolizumab were evaluated: three non-responders and two responders. The demographics and clinical history of the cohort are listed in Supplementary Table 1.

### Spatial Transcriptomic Analysis

A total of 119 Areas of Interest (AOIs) were profiled across five patients at two treatment time points (pre- and post-treatment), capturing 60 tumor (PanCK+) and 59 stromal (PanCK-) segments. As depicted in **Figure 1A**, the application of DSP technology enabled precise delineation, revealing distinct segregation of PanCK and Stromal segments within the tumor microenvironment. Examining differential gene expression between tumoral and stromal segments, we found elevated levels of keratins (KRT7 and KRT19) within the PanCK+ segments. Conversely, PanCK-segments exhibited increased expression of stromal (ACTA2, COL1A1, COL4A1, COL6A1) and immune markers (IGHG4, IGHG2). The enrichment of expected epithelial markers (keratins) in PanCK+ segments, and stromal markers (collagens) in the stromal segments validates the quality of transcriptomic data obtained in this study. Further, this molecular characterization offers valuable insights into the unique profiles of PanCK+ and PanCK-segments, identifying dynamic molecular landscape in BCG unresponsive high-grade NMIBCs **(Figure 1B)**.

**Figure 1:**
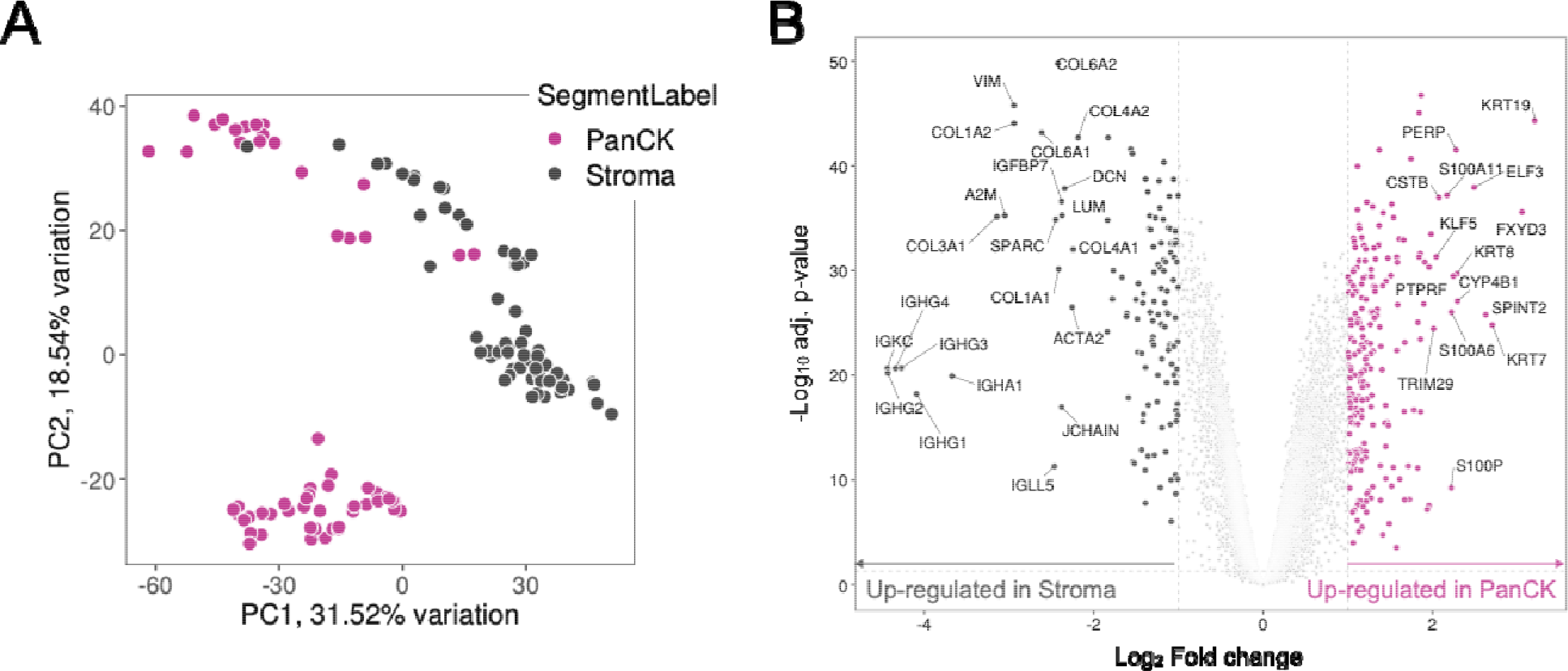
**A)** Principal component analysis visualizing distribution of AOIs in the cohort. **B)** Volcano plot depicting differential expressed genes between PanCK+ tumor and PanCK-stroma segments.

### Characteristics of PanCK+ tumor segments that are predictive of response and Impact of therapy on gene expression in PanCK+ tumor segments

The reported response to pembrolizumab in BCG unresponsive bladder NMIBCs is 40% at three months. To identify expression signatures of the tumor segments that may affect the treatment response, we first characterized the 60 PanCK+ AOIs. Comparing the expression of canonical bladder cancer subtype markers across the cohort **(Figure 2A)**, we identified elevated levels of claudin-low and squamous differentiation markers in pre-treatment PanCK+ segments from responders. In contrast, pre-treatment PanCK+ AOIs from non-responders were enriched for luminal markers. Claudin-low tumors have been previously identified to be immune infiltrated with an expression profile predicted to respond to immune checkpoint blockade^7^.

**Figure 2:**
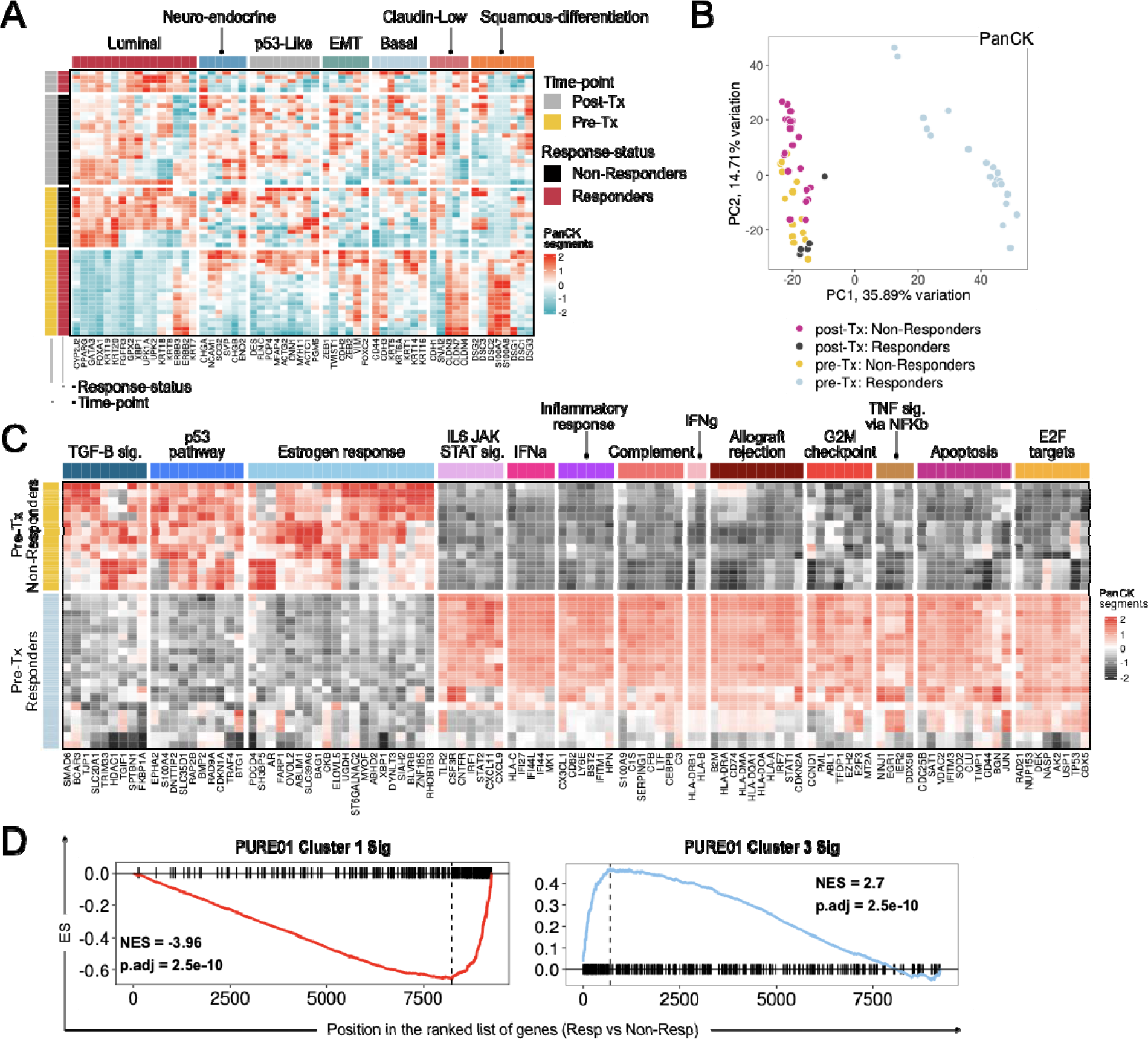
**A)** Heatmap illustrating gene expression patterns across bladder cancer subtypes within PanCK segments. **B)** Principal Component Analysis (PCA) plot showing the distribution and relationships among PanCK segments in the cohort. **C)** Heatmap highlighting genes and pathways significantly enriched in pre-treatment PanCK segments, distinguishing responders from non-responders. **D)** Gene Set Enrichment Analysis (GSEA) plot showcasing the enrichment of PURE01 gene signatures in PanCK segments and comparing responders to non-responders.

We identified minimal heterogeneity of AOIs from each patient. Comparing the distribution of PanCK+ segments, we find that the expression profile of pre-treatment responders formed a distinct cluster independent of the rest of the cohort **(Figure 2B)**. Analysis of pre-treatment PanCK+ AOIs revealed distinctive gene programs associated with treatment response or resistance. Specifically, pre-treatment PanCK+ segments from responders demonstrated elevated inflammation-related pathways, including significant upregulation of genes related to interferon-alpha and gamma response, TNF-alpha signaling, and the IL6-JAK-STAT signaling pathways. **(Figure 2C)**. These findings suggest a heightened inflammatory tumor epithelium within the pre-treatment responder segments. In contrast, PanCK+ segments from non-responders had repressed inflammation pathways. Similar to “immune-excluded” tumors, the tumor epithelium had elevated markers of TGF-beta signaling, p53 pathway genes, and estrogen response. **(Figure 2C)**.

We recently described a transcriptome-based evaluation of the response to immune checkpoint inhibitors in muscle-invasive bladder cancer^6^. We hypothesized that the underlying mechanisms of pembrolizumab activity may remain consistent in early-stage bladder cancer, To test this hypothesis, we applied the five gene signatures described by Robertson et al to tumor AOIs within this cohort^6^. We find Cluster1-MIBC-CPI signatures which were associated with resistance to pembrolizumab (15% complete response) and enriched in luminal immune cold-MIBC tumors with FGFR3 mutations, to be upregulated in pre-treatment PanCK+ segments from non-responders in this cohort. Robertson et al. found Clusters 2 and 3-MIBC-CPI were associated with immune infiltration and a favorable response to immunotherapy (63% complete response)^6^. We find Cluster3 and Cluster2-MIBC-CPI signatures upregulated in pre-treatment PanCK+ segments from responders in the IV pembrolizumab cohort **(Figure 2D and Supp Fig 1A)**. Collectively, this suggests that the CPI clusters may be conserved in the tumor epithelium across stage.

We were interested in the dynamic changes caused by pembrolizumab. Spatial profiling of longitudinally collected specimens pre- and post-therapy allowed us to isolate the impact of therapy on the tumor epithelium and the TME separately. By PCA, PanCK+ AOI segments from non-responders (both pre-and post-treatment) clusters closely, highlighting the minimal impact of treatment on the transcriptomes. The limited change in gene expression diverges from the drastic expression changes found in responsive tumors after treatment **(Figure 2B)**. Specifically, 807 genes were differentially regulated in responsive tumors, compared to only 22 in non-responsive tumors **(Fig 3A)**. To identify the gene sets that change in response to pembrolizumab, we compared PanCK+ segments pre-treatment to those collected post-treatment. As seen in **Figure 3B**, post-treatment PanCK+ segments from responders showed a net increase in inflammation-related pathways. In contrast, post-treatment PanCK+ segments from non-responders had a decrease in inflammatory pathways in response to pembrolizumab, including TNF⍰-signaling, IFN□ response, and IL6-JAK-STAT3 signaling **(Figure 3B)**. Overall, our analysis indicates that PanCK+ segments exhibiting signs of inflammation before IV pembrolizumab administration may display enhanced responsiveness to this therapy.

**Figure 3:**
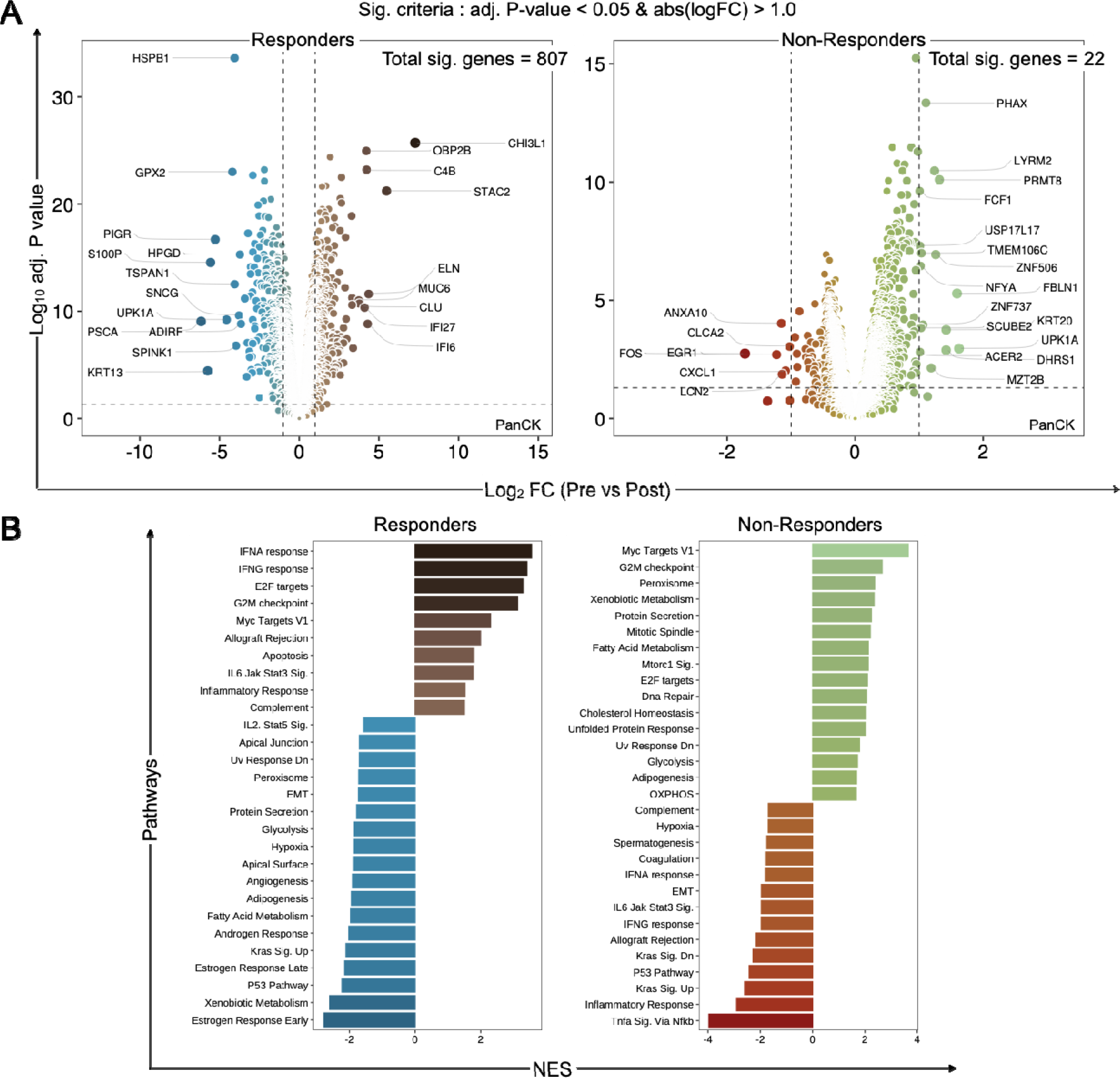
**A)** Volcano plot comparing gene expression profiles between pre-treatment and post-treatment PanCK+ segments from responders and non-responders. **B)** Bar plot highlighting pathways changing in responders and non-responders pre-to-post treatment.

### Characteristics of PanCK-stromal segments that are predictive of response

We next profiled the features of the tumor microenvironment (TME) that could contribute to responsiveness to pembrolizumab. As seen in **Figure 4A**, the pre-treatment TME areas of interest (AOIs) from responders cluster separately from non-responders. Comparing the gene expression programs, we find inflammatory markers such as IFN⍰and IFN□ strongly upregulated in the pre-treatment stromal AOIs from responders. In comparison, non-responder PanCK-AOIs had upregulation of EMT, myogenesis, angiogenesis, and coagulation gene sets **(Figure 4B)**. We utilized immune deconvolution to identify individual immune cell populations within each stromal AOI. TMEs from responders had elevated levels of neutrophils, T cells, and NK cells in the tumor microenvironment relative to non-responders **(Figure 4C)**. In response to treatment, we observed an increase in the myeloid cell population in responders and a decrease in non-responders **(Figure 4C)**.

**Figure 4:**
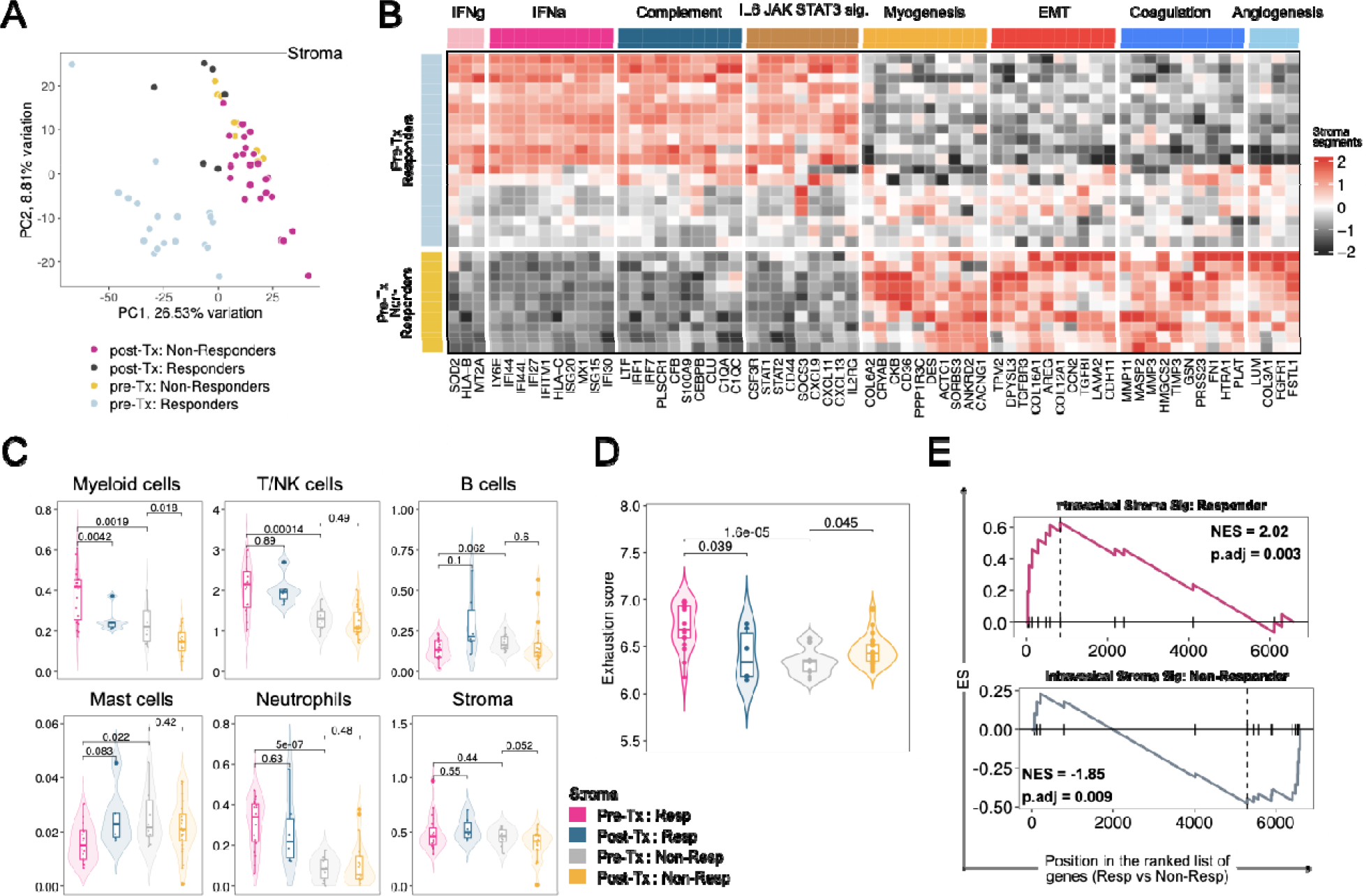
**A)** Principal component analysis visualizing distribution of Stromal AOIs in the cohort. **B)** Heatmap highlighting pathways significantly enriched in pre-treatment stromal segments from responders and non-responders. **C)** Violin boxplots comparing cellular abundance of specified immune populations within the indicated conditions. **D)** Violin boxplots comparing exhaustion scores between responders and non-responders in both pre-and post-treatment samples. **E)** GSEA plot showing the enrichment of stromal gene signatures generated in the intravesical cohort in differentially expressed genes from stromal segments from the IV pembrolizumab cohort comparing responders to non-responders.

Pembrolizumab is an immune checkpoint blocking antibody and its efficacy partly relies on the expression of PD-1 on the immune cell surface. Overexpression of PD-1 on the cell surface is a well-established marker of T-cell exhaustion. To understand the functional state of immune cells inhabiting the stromal compartment, we compared expression of exhaustion markers across the different conditions and identified elevated levels of this gene set in the pre-treatment stromal compartment from responders relative to non-responders **(Figure 4D)**. Interestingly, we found a minor but significant decrease in the exhaustion score in responders post-treatment. However, this contrasted with an increased exhaustion score observed in non-responders **(Figure 4D)**. Previous work from our group has focused on studying tumor-immune dynamics associated with response to intravesical BCG and pembrolizumab combination therapy^3^. We applied stromal gene signatures that predicts response or resistance to BCG and pembrolizumab and we were able to validate these two pre-treatment signatures in tumors treated IV pembrolizumab. **(Figure 4E)**.

### Comparison of response strategies for high-risk non-muscle invasive bladder cancer

Using the same platform, our group previously described the spatial comparisons of the first-in-human administration of BCG and intravesical pembrolizumab^3^. Given this cohort’s unique administration of pembrolizumab, we wanted to evaluate the differences in urothelial gene expression profile between intravesical pembrolizumab and BCG compared IV pembrolizumab. One of the significant benefits of the Digital Spatial Profiling platform is the ability to assay the tumor and stroma segments in physical proximity. We measured if the correlation between the inflammation levels of the PanCK+ segments and the immune infiltration levels of the neighboring stromal segments in pre-treatment AOIs are different between responders and non-responders between the two cohorts. As seen in **Figure 5A**, responders to the combination intravesical therapy exhibited low levels of inflammation in the PanCK+ segments and high levels of infiltration in the neighboring stromal segments. However, responders to the IV pembrolizumab monotherapy showed high levels of inflammation in the pre-treatment PanCK+ segments and high levels of immune infiltration in the adjacent stroma.

**Figure 5:**
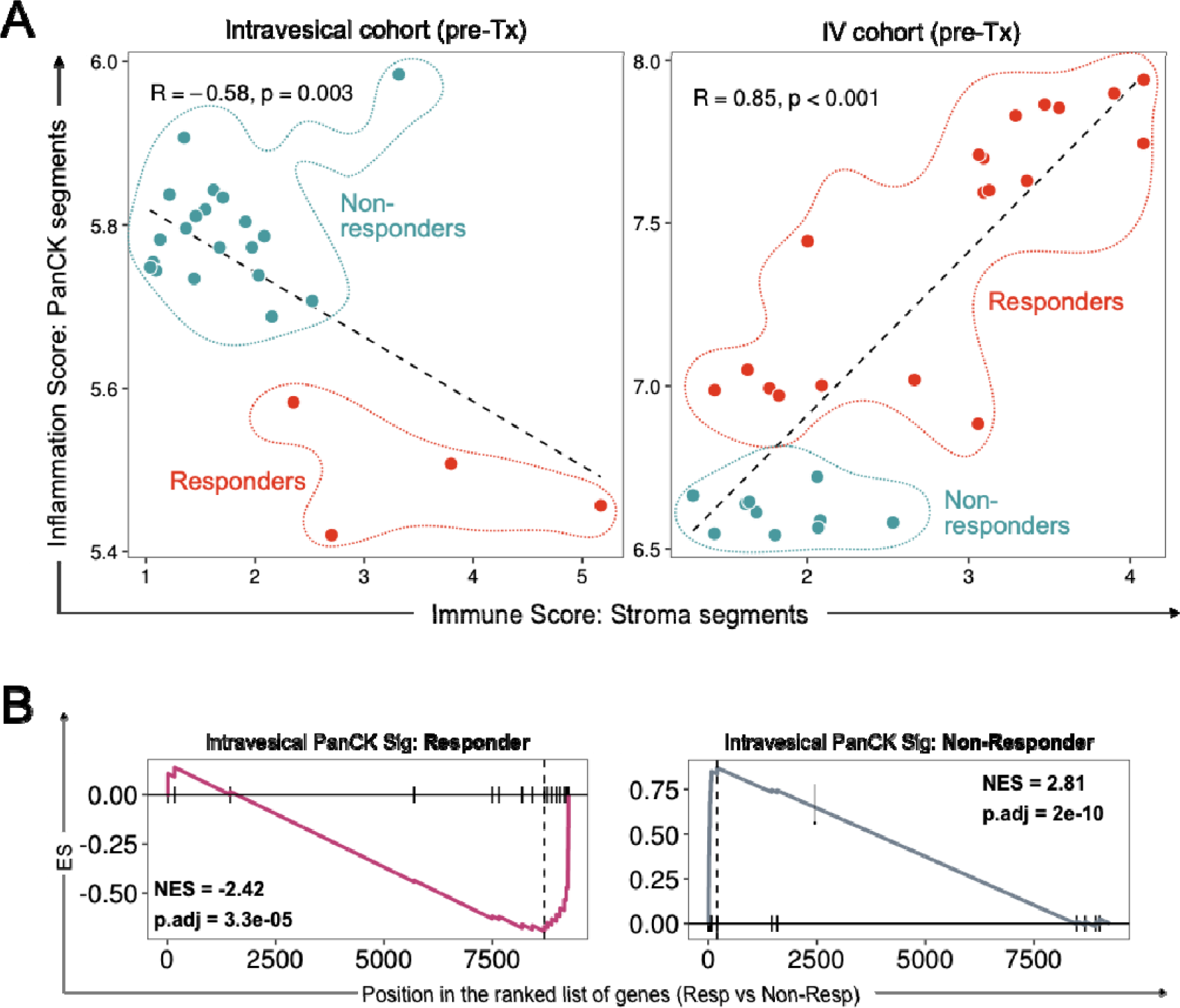
**A)** GSEA plot showing the enrichment of stromal gene signatures generated in the intravesical cohort in differentially expressed genes from stromal segments from the IV pembrolizumab cohort comparing responders to non-responders. **B)** Correlation plot comparing inflammation score for PanCK segments and infiltration score in the neighboring Stromal segments between responders and non-responders for the intravesical BCG+Pembrolizumab and IV Pembrolizumab Cohort.

Consistent with these differences, we tested the PanCK+ signatures generated in the intravesical study to determine if they can assess response in the tumors treated with IV pembrolizumab. As anticipated, we find that the PanCK+ signature that predicts response in the intravesical pembrolizumab and BCG cohort identified non-responders in the pembrolizumab IV cohort, and the PanCK+ signature that predicts lack of response in the intravesical combination therapy cohort are enriched in responders in the pembrolizumab monotherapy cohort **(Figure 5B)**.

This comparison suggests that response to IV pembrolizumab is related to inflammation in the tumor and TME. Overall, our findings suggest tumor segments that are inflamed before therapy might benefit more from IV pembrolizumab, whereas pre-treatment non-inflamed BCG unresponsive tumors might be a better candidate for treatment with a combination therapy of BCG and pembrolizumab.

## Discussion

From the time of the initial reports on immune checkpoint inhibitor efficacy, immunotherapy has been utilized to manage bladder cancers from early to metastatic stages^8^. Overall, the response rate to CPIs in metastatic urothelial cancer is ∼20%, with patients expressing PDL1/PD1 more likely to have a more durable response, a stratified HR 0.68, 95% CI 0.43–1.08 compared to chemotherapy^9,10^. In early-stage BCG unresponsive NMIBC, a similar response was described for patients with CIS, with or without papillary tumors in KEYNOTE057^1,2^ and SWOG S1605^11^. These results suggest similar drug activity and provide a rationale for identifying the specific patients who might benefit long-term from CPI. In this report, we attempt to profile early-stage NMIBCs to identify response mechanisms to CPI. Despite the frequency of BCG unresponsive BCa, few patients are cured (NED for > 24 months). Yet, the tail of response appears stable for <20% of patients with pembrolizumab. Our goal with this investigation was to identify pre-treatment features associated with response or lack thereof.

We evaluated the tumor and TME of responders and non-responders and identified pre-treatment immune signatures associated with response to IV pembrolizumab. We validated signatures generated from PURE01-CPI trial and the intravesical BCG and pembrolizumab trial. Our data confirms that resistance to pembrolizumab is a consequence of limited immune infiltration into the tumor microenvironment^3,6,12^. It is notable that the pre-treatment signature predictive of response to IV pembrolizumab in BCG unresponsive disease seen here (inflamed tumor epithelium and infiltrated stroma) differs from our previous work with combination intravesical BCG and pembrolizumab (non-inflamed tumors had better response). Given the sequences of therapy in these two studies (BCG failure followed by IV pembrolizumab vs BCG failure followed by simultaneous administration of intravesical BCG and pembrolizumab), it is possible that most of the benefit from BCG involves inducing an inflammatory anti-tumor response where none is present, while pembrolizumab ‘releases the breaks’ on an already present yet ineffective inflammatory response from previous BCG. Lastly, our results suggest that the immune response to pembrolizumab is conserved across bladder cancer stages.

Our findings strongly highlight the need to assay the transcriptomic state of the BCG unresponsive tumors prior to deciding the course of treatment. We anticipate that the application of an expression-based biomarker such as the one described here has the potential to identify tumors that are likely to respond to pembrolizumab. Further evaluation of more patients treated with different CPIs is needed to refine our results.

## Conclusion

We performed a spatial-based evaluation of tumors treated with pembrolizumab. We identified distinct expression signatures associated with the response and resistance of the tumor and TME. Future studies evaluating the accuracy of these signatures will help validate our findings and facilitate biomarker application in patients with NMIBC.

## Data Availability

All data produced in the present study are available upon reasonable request to the authors

## Acknowledgments

None

