## Supplementary material for "A Spatial Comparison of Molecular Features Associated with Resistance to Pembrolizumab in BCG Unresponsive Bladder Cancer": Supp Fig 1

**Supp Figure 1:**

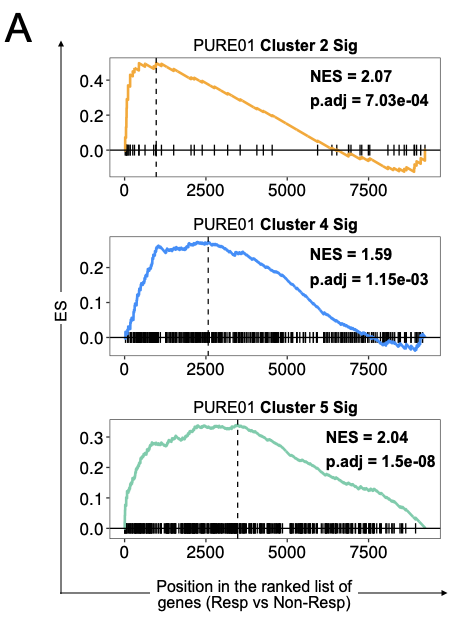

**Supp Figure 1:**

A) GSEA plot highlighting enrichment of indicated PURE01 gene signatures in PanCK segments comparing responders vs non-responders.
