## Supplementary material for "A Spatial Comparison of Molecular Features Associated with Resistance to Pembrolizumab in BCG Unresponsive Bladder Cancer": Supp Table 1

**Supp Table 1:** Patient Demographics and Oncologic History

Demographic and oncologic outcomes of patients included in the study. The listed age is at the time of collection for the first specimen. Pre- and post-treatment pathology lists pathologic stage for specimens used for digital spatial profiling.

| **Patient #** | **Age** | **Sex** | **Pre-treatment pathology** | **Post-treatment pathology** | **Response** | **Smoking Status** | **Pembrolizumab Doses** | **Time on Therapy (days)** | **Outcome** |
| --- | --- | --- | --- | --- | --- | --- | --- | --- | --- |
| 1 | 75-80 | M | Tis | Tis | NR | Non-smoker | 6 | 109 | Progression to MIBC, T4aN0 at cystectomy |
| 2 | 75-80 | F | TaHG/CIS | T1HG/CIS | NR | Former smoker (>20 pack years) | 22 | 675 | Progression to metastatic disease |
| 3 | 65-70 | M | Tis | T0 | R | Non-smoker | 20 | 1179 | Remains NED |
| 4 | 75-80 | M | T1HG | Tis (minimal) | R | Former smoker (>20 pack years) | 11 | 285 | Minimal persistent CIS, completely resected - now NED |
| 5 | 85-90 | M | TaHG/CIS | Tis (diffuse) | NR | Former smoker (>20 pack years) | 5 | 87 | Persistent diffuse CIS |

**Abbreviations** : BCG: Bacillus Calmette-guérin, CIS: Carcinoma in situ, HG: High-Grade, MIBC: Muscle Invasive Bladder Cancer NED: No Evidence of Disease, NR: Non-responder to pembrolizumab, R: Responder to pembrolizumab UCC: Urothelial Cell Carcinoma.
